# Knowledge and attitudes towards dementia care among healthcare workers in Somaliland: a need assessment survey

**DOI:** 10.1101/2023.10.17.23297149

**Authors:** Mohamed Abdilahi Duale, Tewelde Tesfaye Gebremariam

## Abstract

The present pilot survey assessed the levels of healthcare workers’ knowledge of dementia and attitudes towards dementia care in Somaliland. Between June and July 2023, we administered an adapted online survey with 30 Alzheimer’s disease Knowledge Scale (ADKS) true or false statements and 10 Dementia Care Attitude Scale (DCAS) Likert-scale items. The reliability of the Likert-scale DCAS items was assessed using Cronbach’s alpha. A Chi-square, Mann-Whitney U-test, and independent sample t-test were used to assess the relationship and differences of the outcome variables (knowledge and attitude) across the healthcare workers’ baseline information. The mean ADKS score was 18.7 (SD = 3.1). Out of 107 healthcare workers who completed the survey, 49.5% had high dementia knowledge (score ≥18.7). The participants’ median (IQR) attitude score was 34 (25, 39), and 46.7% (50 of 107) had a positive attitude towards dementia care. In conclusion, health professionals in Somaliland had low levels of knowledge and negative attitudes towards dementia care.

## Introduction

Dementia, mainly Alzheimer’s disease (AD), is the seventh leading cause of disability, dependency, and death among older people globally. More than 55 million individuals worldwide suffer from the disease, with more than 60% living in low- and middle-income nations (LMICs). Dementia disproportionately affects older people over 65 and women [1]. Healthcare workers, mainly primary physicians and community nurses, are key players in providing dementia care by counseling, connecting with, and helping older adults with dementia [2, 3].

The Republic of Somaliland is a self-declared young country in the Horn of Africa. The nation has experienced recurrent and devastating droughts and extreme hunger since its independence in 1991 [4]. It is home to 5.7 million people, and older people over 65 years-old account for 4% of the total population [5, 6]. Due to other competing health priorities in Somaliland, elderly people are often forgotten, and their specific healthcare needs remain largely ignored. For instance, dementia palliative care education and training are not included in the curricula for health professional students or continuing professional development programs. Hence, identifying evidence to inform dementia care education and training requires assessing health workers’ knowledge and attitudes concerning dementia and dementia care.

Our pilot needs assessment survey, the first of its kind in the Republic of Somaliland, aimed to assess health professionals’ dementia knowledge and attitudes towards dementia care.

## Materials and methods

### Study design and setting

Between June and July 2023, we undertook an online pilot survey with a convenient sample of 107 health professionals in Somaliland. About 197 doctors, 1256 nurses, and 344 midwives serve the country’s population [7]. English is the official academic language used in higher education in the self-declared country.

### Data collection tool development and measurement

We used an English questionnaire with Alzheimer’s Disease Knowledge Scale (ADKS) statements [8] and Dementia Care Attitude Scale (DCAS) statements [9] adapted from previous studies. We assessed the health professionals’ knowledge about Alzheimer’s disease and attitudes towards dementia with ADKS and DCAS, respectively. The ADKS is a single-factor scale that includes 30 true or false questions covering seven domains: life impact (3 items), assessment and diagnosis (4 items), symptoms (4 items), course of the disease (4 items), treatment and management (4 items), caregiving (5 items), and risk factors (6 items) [2]. We determined the total score by adding the correct responses for each item, resulting in a total score ranging from 0 to 30. A higher overall score implies better knowledge [9]. The mean ADKS score was used as a benchmark to determine the knowledge level of the participants. We regarded the knowledge level as high if the score was equal to or higher than the mean score; otherwise, we deemed it low.

The DCAS contains 10 items, with responses scored on a 5-point Likert scale ranging from 1 (strongly disagree) to 5 (strongly agree). Four of the 10 items are negatively worded and are reversed in a definite order (e.g., a score of 5 becomes 1) when calculating the total score. The total score range was from 10 to 50; scores above the median attitude score were considered positive attitudes, and scores below the median score were considered negative attitudes.

### Recruitment process

Directors and principals of health institutions were requested to circulate the survey link to healthcare workers in the Republic of Somaliland, who in turn were encouraged to further share the link with other participants in health facilities across the country.

### Survey administration

Kobo Toolbox forms were administered (see also the supplementary file), and participation was voluntary, and no incentives were offered. We limited participation to a single response.

## Data management and analysis

We exported and downloaded the Kobo Toolbox data in XLS forms and cleaned and coded it in Excel before importing it into SPSS. We used Cronbach’s alpha coefficient [10] to assess the reliability of the attitude statements. We used the Kolmogorov-Smirnov test to assess the normality of all the scale variables.

We summarized baseline information (such as age, gender, and education level) and metrics of main outcome data (ADKS and DCAS) using descriptive statistics, such as frequency, percentage, and measures of central tendency (mean and median) and measures of dispersion (standard deviation and interquartile range).

We performed the chi-square test to investigate the relationship between baseline characteristics and ADKS and DCAS, the Mann-Whitney U test to assess the difference in DCAS score, and the independent sample t-test to assess the difference in ADKS score across the baseline characteristics of the participants. All tests were performed using SPSS v. 20 (Armonk, NY, IBM, Corp.) [11], and the statistical significance was set at a two-tail *p*-value < 0.05.

### Ethics statement

Ethical approval was obtained from the Frantz Fanon Institutional Review Board (FFU/IRB/00000001) (see supplementary file). We collected no personally identifiable information. On the first page of the online questionnaire, all relevant information regarding the survey, data protection, and the statement of consent was presented. Participants provided informed consent before undertaking this survey, and permission was granted for publication of the results.

## Results

### Baseline characteristics

Out of the 107 healthcare workers who completed the online survey, more than half were aged between 18 and 39 years old (97.2%), had bachelor’s and above (91.8%), were men (51.4%), and had experience caring for people with dementia (62.6%) (Table **1**).

**Table 1.** Summary of participants’ baseline characteristics.

| **Characteristics** | **Frequency** | **Percentage** |
| --- | --- | --- |
| Age, median (IQR) | 28(25,31) | 25,31 |
| 18-39 | 103 | 97.2 |
| 40-59 | 2 | 1.9 |
| ≥60 | 1 | 0.9 |
| Gender |  |  |
| Male | 55 | 51.4 |
| Female | 52 | 48.6 |
| Education Level |  |  |
| Diploma | 8 | 8.2 |
| Bachelor and above | 90 | 91.8 |
| Experience in dementia care |  |  |
| No | 40 | 37.4 |
| Yes | 67 | 62.6 |

### Alzheimer’s disease Knowledge Scale (ADKS)

The Alzheimer’s Disease Knowledge Scale (ADKS) mean score was 18.7 (SD = 3.1) out of 30, and healthcare workers responded to 62% of the ADKS statements correctly (Table **2**, Table **3**).

**Table 2.** Participants’ Alzheimer’s Disease Knowledge Disease Scale (ADKS).

| ADKS Domains | Answered correctly<br>N (%) | Mean ± SD |
| --- | --- | --- |
| **Life Impact** |  |  |
| It is safe for people with Alzheimer's disease (AD) to drive as long as they have a companion in the car at all times. (F) | 62 (57.9) | 0.58 ± 0.50 |
| Most people with AD live in nursing homes. (F) | 56 (52.3) | 0.52 ± 0.50 |
| Drivers in the early stages of AD have more auto accidents than other older drivers. (F) | 21 (19.6) | 0.20 ± 0.40 |
| **Domain mean score ± SD** |  | **1.30 ± 0.88** |
| **Assessment and Diagnosis** |  |  |
| An evaluation of a person for AD typically includes information from a physical exam, memory tests, brain scans, and a history of the symptoms. (T) | 98 (91.6) | 0.92 ± 0.23 |
| AD is a normal part of aging, like gray hair or wrinkles. (F) | 60 (56.1) | 0.56 ± 0.50 |
| Currently, the best way to diagnose AD is with a blood test. (T) | 33 (30.8) | 0.31 ± 0.46 |
| A person suspected of having AD should be evaluated to rule out treatable disorders with similar symptoms. (T) | 94 (87.9) | 0.88 ± 0.33 |
| **Domain mean score ± SD** |  | **2.63 ± 0.97** |
| **Symptoms** |  |  |
| AD progresses at the same speed as everyone else. (F) | 79 (73.8) | 0.74 ± 0.44 |
| In general, as people with AD get worse, they are likely to wander and get lost. (T) | 90 (84.1) | 0.84 ± 0.37 |
| Some people with AD cannot recognize their children when they see them. (T) | 89 (83.2) | 0.83 ± 0.38 |
| People with AD have more problems remembering things on some days than others. (T) | 87 (81.3) | 0.74 ± 0.39 |
| **Domain mean score ± SD** |  | **3.22 ± 0.90** |
| **The course of the disease** |  |  |
| In rare cases, people have recovered from Alzheimer's disease. (F) | 42 (39.3) | 0.39 ± 0.49 |
| Eventually, the person with AD will need 24-hour supervision. (T) | 87 (81.3) | 0.81 ± 0.39 |
| After symptoms of AD appear, the average life expectancy is 6-12 years. (T) | 62 (57.9) | 0.58 ± 0.50 |
| A person with AD becomes increasingly likely to fall as the disease gets worse. (T) | 87 (81.3) | 0.81 ± 0.40 |
| **Domain mean score ± SD** |  | **2.53 ± 1.01** |
| **Treatment and Management** |  |  |
| AD cannot be cured. (T) | 80 (74.8) | 0.75 ± 0.44 |
| When a person has AD, using reminder notes is a crutch that contributes to decline. (F) | 40 (37.4) | 0.37 ± 0.49 |
| People whose AD is not severe can benefit from psychotherapy for depression and anxiety. (T) | 89 (83.2) | 0.83 ± 0.38 |
| Medications can permanently stop AD from getting worse. (F) | 50 (46.7) | 0.47 ± 0.50 |
| **Domain mean score ± SD** |  | **2.42 ± 1.00** |
| **Caregiving** |  |  |
| People with AD do best with simple instructions given one step at a time. (T) | 83 (77.6) | 0.78 ± 0.42 |
| When people with AD repeat the same question or story several times, it is helpful to remind them that they are repeating themselves. (F) | 33 (30.8) | 0.31 ± 0.46 |
| If a person with AD becomes alert and agitated at night, a good strategy is to make sure that the person gets plenty of physical activity during the day. (T) | 79 (73.8) | 0.74 ± 0.44 |
| If a person with AD follows the caregiver all over the house, it is helpful to encourage the person with AD to stay in one room. (F) | 44 (41.1) | 0.41 ± 0.50 |
| People with AD do best when exposed to new experiences and environments as often as possible. (F) | 37 (34.6) | 0.35 ± 0.50 |
| **Domain mean score ± SD** |  | **2.58 ± 1.00** |
| **Risk factors** |  |  |
| Genes can only partially account for the development of dementia. (T) | 72 (67.3) | 0.67 ± 0.47 |
| AD can be caused by eating food that is cooked in aluminum pots. (F) | 76 (71.0) | 0.71 ± 0.50 |
| More than 50% of people older than 85 have AD. (T) | 83 (77.6) | 0.78 ± 0.42 |
| The percentage of people older than 65 with AD exceeds 10%. (F) | 26 (24.3) | 0.24 ± 0.43 |
| Having a parent or sibling with AD increases the chance of developing it. (T) | 82 (76.6) | 0.77 ± 0.43 |
| It has been scientifically proven that mental exercise can prevent a person from getting AD. (T) | 80 (74.8) | 0.75 ± 0.44 |
| **Domain mean score ± SD** |  | **3.92 ± 1.07** |
| **Overall ADKS mean score ± SD** |  | **18.70±3.13** |

**Table 3.** Summary of healthcare workers’ responses across the Alzheimer’s disease Knowledge Scale (ADKS) domains.

| ADKS domains | # of items | %Correct responses | Mean ±SD |
| --- | --- | --- | --- |
| Life impact | 3 | 43% | 1.30 ± 0.88 |
| Assessment and diagnosis | 4 | 66% | 2.63 ± 0.97 |
| Symptoms | 4 | 80% | 3.22 ± 0.90 |
| Course of the disease | 4 | 66% | 2.53 ± 1.01 |
| Treatment and management | 4 | 61% | 2.42 ± 1.00 |
| Caregiving | 5 | 52% | 2.58 ± 1.00 |
| Risk factors | 6 | 65% | 3.92 ± 1.07 |
| **Overall** | **30** | **62%** | **18.70±3.13** |

Items with the poorest responses included those related to life impact and caregiving, with 43% and 52% correct answers, respectively (Table **3**). More than half of participants mistakenly thought that drivers in the early stages of Alzheimer’s disease (AD) have more auto accidents than other older drivers (80%, n = 86); when people with AD repeat the same question or story several times, it is helpful to remind them that they are repeating themselves (69%, n = 74); and people with AD do best when exposed to new experiences and environments as often as possible (65%, n = 70) (Table **2**).

Furthermore, a 61% correct response rate was recorded about dementia treatment and management, and about 37% and 47% responded correctly that ‘when a person has AD, using reminder notes is a crutch that contributes to decline’ and ‘Medications can permanently stop AD from getting worse’, respectively (Table **2**).

Out of the 107 participants, 49.5% had a high knowledge level (ADKS score ≥18.7) (Fig. **1**). The knowledge level was significantly associated with gender (χ^2^ = 4.11, p-value = 0.043), and women had a higher mean ADKS score than men (Fig. **2**, Fig. **3**, t-test = -2.441, *p* = 0.016).

**Fig. 1.**
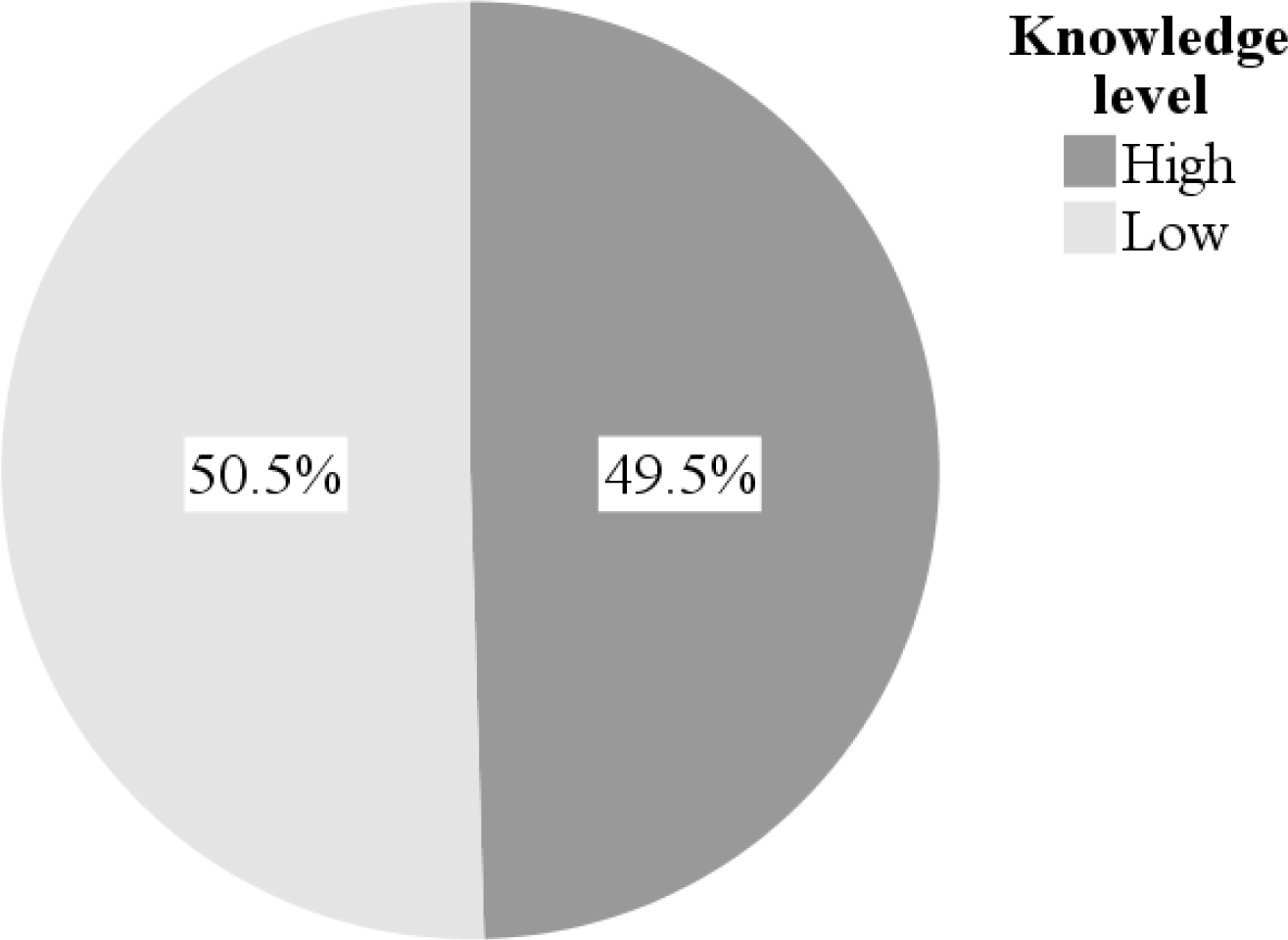
Knowledge level of participants on Alzheimer’s disease.

**Fig. 2.**
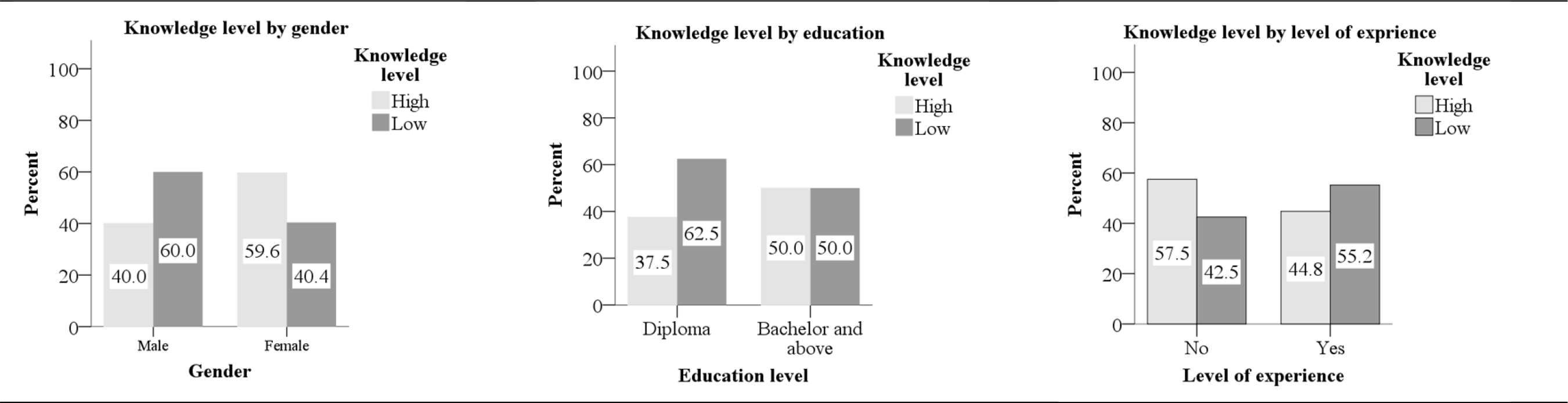
Knowledge level on Alzheimer disease by baseline characteristics.

**Fig. 3.**
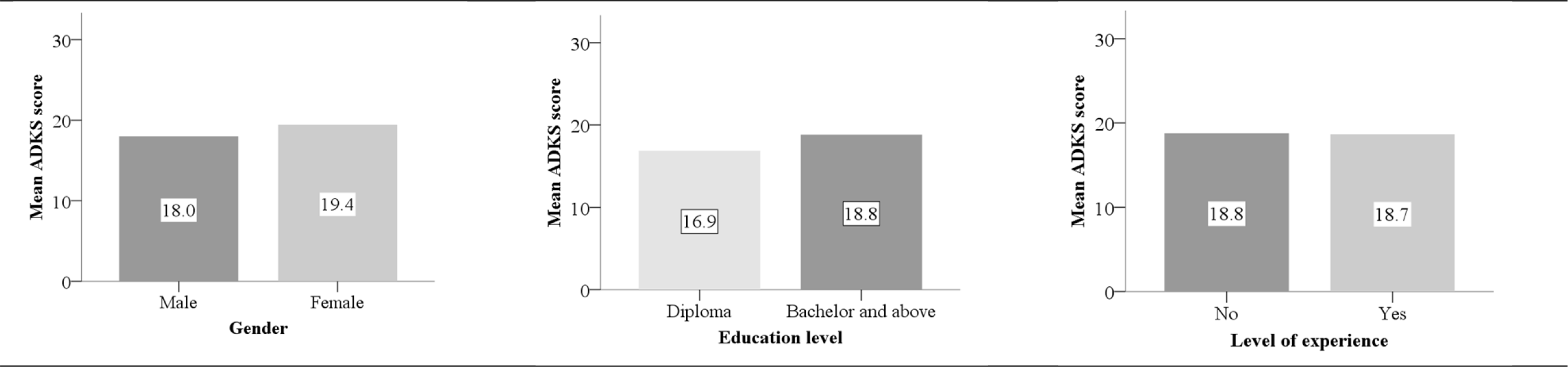
Mean ADKS score by baseline characteristics.

### Dementia Care Attitude Scale (DCAS)

The study showed that the DCAS questionnaire demonstrated adequate internal consistency; the overall reliability (measured as Cronbach’s alpha) value of the scale was 0.88, while the item values ranged from 0.86 to 0.89. A two-way mixed effect model and average consistency measure were used to measure the interclass correlation of the items, which was 0.88 (95% CI 0.85, 0.91).

The median (IQR) DCAS score was 34 (25, 39) (Table **4**), and 46.7% of participants (50 of 107) had a positive attitude towards dementia care (DCAS score ≥34) (Fig. **5**). Over three-quarters of healthcare workers mistakenly disapproved that providing diagnosis is usually more helpful than harmful (Table **4**, Fig. **4**). Likewise, over one-third of participants had a negative view that ‘managing dementia is more often frustrating than rewarding’, ‘there is little point in referring families to services as they do not want to use them’, ‘the primary care team has very little role to play in the care of people with dementia’, and ‘patients with dementia can be a drain on resources with little positive outcome’ (Table **4**, Fig. **4**).

**Table 4.** Distribution participants’ responses to dementia care attitude scale (DCAS) statements.

| DCAS statements | Strongly agree | Agree | Neutral | Disagree | Strongly disagree | Mean ± SD |
| --- | --- | --- | --- | --- | --- | --- |
| Much can be done to improve the quality of life of caregivers of people with dementia. (PW) | 38 (35.5%) | 26 (24.3%) | 9 (8.4%) | 10 (9.3%) | 24 (22.4%) | 3.41±1.58 |
| Families would rather be told about their relatives' dementia as soon as possible. (PW) | 24 (22.4%) | 37 (34.6%) | 11 (10.3%) | 9 (8.4%) | 26 (24.3%) | 3.22±1.51 |
| Much can be done to improve the quality of life of people with dementia. (PW) | 33 (30.8%) | 33 (30.8%) | 12 (11.2%) | 10 (9.3%) | 19 (17.8%) | 3.48±1.46 |
| Providing a diagnosis is usually more helpful than harmful. (PW) | 38 (35.5%) | 37 (34.6%) | 9 (8.4%) | 3 (2.8%) | 20 (18.7%) | 3.65±1.46 |
| Dementia is best diagnosed by specialist services. (PW) | 27 (25.2%) | 35 (32.7%) | 13 (12.1%) | 12 (11.2%) | 20 (18.7%) | 3.35±1.45 |
| Patients with dementia can be a drain on resources with few positive outcomes. (NW) | 6 (5.6%) | 29 (27.1%) | 30 (28.0%) | 19 (17.8%) | 23 (21.5%) | 2.78±1.22 |
| It is better to talk to the patient in euphemistic terms. (PW) | 16 (15.0%) | 30 (28.0%) | 30 (28.0%) | 14 (13.1%) | 17 (15.9%) | 3.13±1.28 |
| Managing dementia is more often frustrating than rewarding. (NW) | 12 (11.2%) | 27 (5.2%) | 24 (22.4%) | 19 (17.8%) | 25 (23.4%) | 2.83±1.34 |
| There is little point in referring families to services as they do not want to use them. (NW) | 6 (5.6%) | 30 (28.0%) | 31 (29.0%) | 20 (18.7%) | 20 (18.7%) | 2.83±1.19 |
| The primary care team has very little role to play in the care of people with dementia. (NW) | 24 (22.4%) | 19 (17.8%) | 13 (12.1%) | 18 (16.8%) | 33 (30.8%) | 2.84±1.57 |
| **Overall median (IQR) attitude score** |  |  |  |  |  | **34(25, 39)** |

**Fig. 4.**
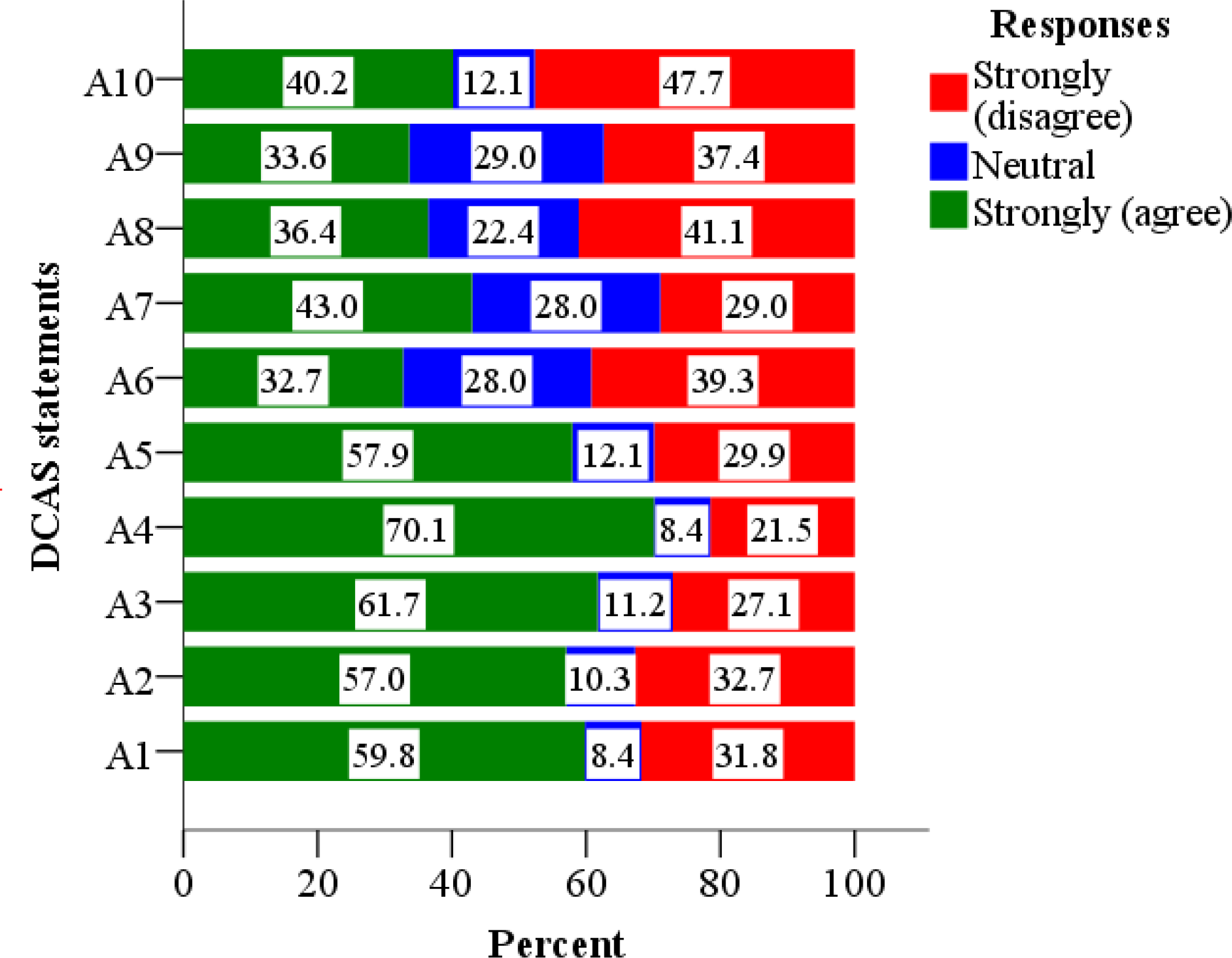
Stack chart showing distribution of healthcare workers’ response to Dementia Care Attitude Scale statements. **A1**: Much can be done to improve the quality of life of caregivers of people with dementia; **A2**: Families would rather be told about their relatives’ dementia as soon as possible; **A3**: Much can be done to improve the quality of life of people with dementia; **A4**: Providing a diagnosis is usually more helpful than harmful; **A5**: Dementia is best diagnosed by specialist services; **A6**: Patients with dementia can be a drain on resources with little positive outcomes; **A7**: It is better to talk to the patient in euphemistic terms; **A8**: Managing dementia is more often frustrating than rewarding; **A9**: There is little point in referring families to services as they do not want to use them; **A10**: The primary care team has very little role to play in the care of people with dementia.

**Fig. 5.**
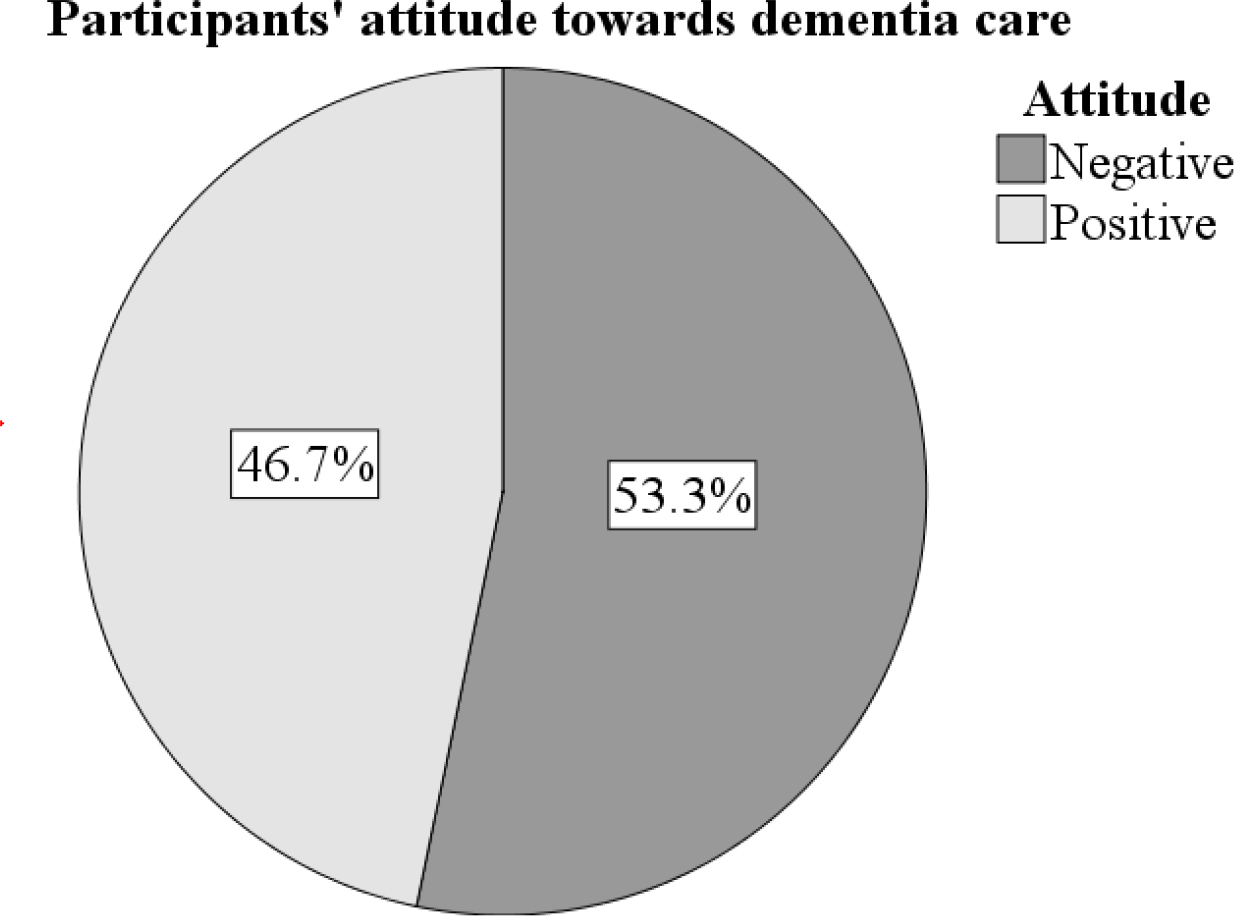
Healthcare workers’ attitude towards dementia care, Somaliland, 2023.

The attitude status was significantly associated with experience in dementia care (χ^2^ = 6.38, p-value = 0.012); however, no significant difference in DCAS score or attitude status was documented between those who had experience and those who had not (Fig. **6**, **Error! Reference source not found**., Mann-Whitney U = 1,908.5, *p* = 0.119).

**Fig. 6.**
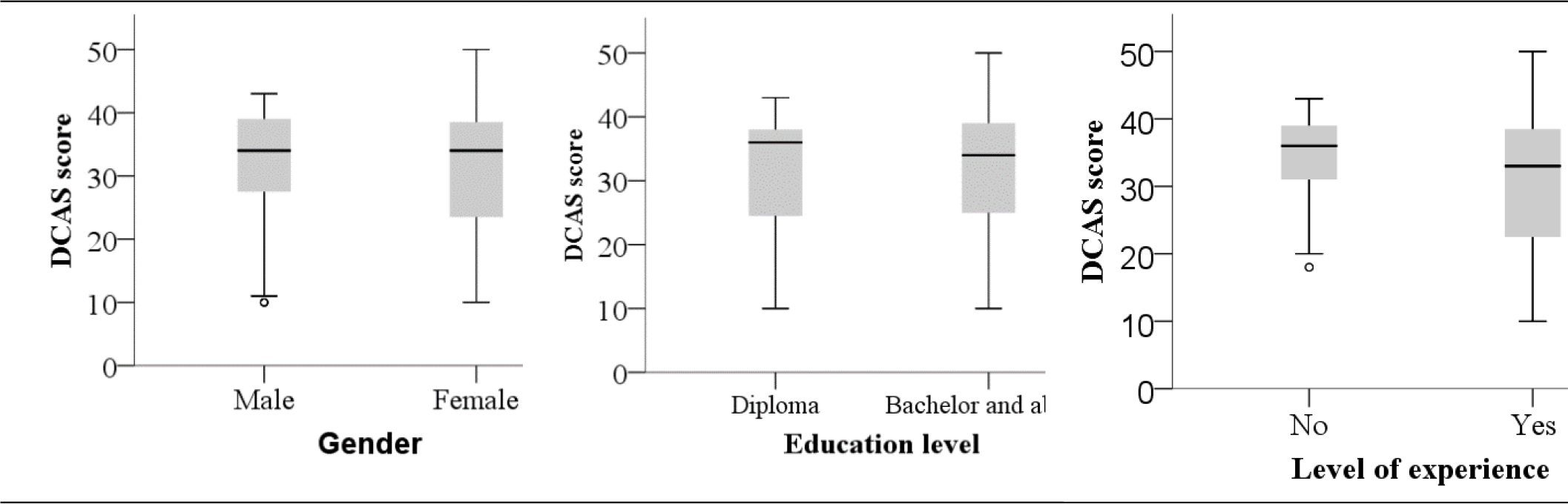
Dementia Care Attitude Scale (DCAS) median (IQR) score by baseline characteristics of participants.

## Discussion

Our pilot study, the first of its kind, assessed healthcare workers’ knowledge of dementia and attitudes towards dementia care in the Republic of Somaliland. Our findings revealed that healthcare providers in Somaliland had low levels of dementia knowledge and attitudes towards dementia care.

The mean Alzheimer’s’ Disease Knowledge Scale (ADKS) score was 18.7, and half (49.5%) of the healthcare workers had a high knowledge level on dementia, which is lower than those reported in Uganda [12], China [2, 9], and Australia [13]. The lowest-level knowledge was related to dementia’s life impact and caregiving. These findings also support previous studies of health professionals in China [2] and Norway [14]. Over half the healthcare workers agreed with incorrect statements that ‘when people with AD repeat the same question or story several times, it is helpful to remind them that they are repeating themselves’, ‘if a person with AD follows the caregiver all over the house, it is helpful to encourage the person with AD to stay in one room’, and ‘people with AD do best when exposed to new experiences and environments as often as possible’. This implies that healthcare workers need to be knowledgeable about caregiving to help people with dementia maintain their autonomy and independence.

Our survey also showed that more than half of the healthcare workers had negative attitudes towards dementia care, and the overall median (IQR) attitude score was 34 (25, 39), which was lower than those reported in China [9].

In conclusion, our need assessment survey demonstrated that healthcare workers in Somaliland had low levels of knowledge and negative attitudes towards dementia. This urges the higher education institutions to embed dementia education into curricula for health professional students or continuing professional development programs. Moreover, a large-scale survey is needed to confirm the above conclusions.

## Acknowledgement

The authors would like to thank everyone who participated in this survey.

## Authors’ contributions

Mr. Gebremariam conceived, designed the study, and prepared the data collection tools. Both Mr. Gebremariam and Mr. Duale conducted the data collection, performed data analysis, and wrote the first draft of the manuscript. Both authors read and approved the final manuscript.

## Funding

No funding was received for conducting this survey.

## Data availability

Data that supports the findings would be available at the request of the corresponding author.

## Code availability

N/A

## Conflict of interest

The authors have no conflict of interest to declare.

## Notes

### Competing Interest Statement

The authors have declared no competing interest.

### Funding Statement

The author(s) received no specific funding for this work.

### Author Declarations

Frantz Fanon University Institutional Review Board approved the protocol (FFU/IRB/00000001).

